# A Cross Sectional Study about The Effects of Smoking on Sleep Apnea in a Sample of Syrian Society

**DOI:** 10.1101/2024.02.16.24302743

**Authors:** Deena al chaar, Mudar al chaar, Hussam Bardan, Adele al chaar

## Abstract

**Background:** To explore whether the use of cigarettes affects the prevalence of obstructive sleep apnea (OSA) in adults.

**Methods:** A questionnaire-based cross-sectional study was conducted among 233 participants. The questionnaire link was published on November 13, 2023 on social media sites and the Internet.The adults were divided into four groups: noncurrent smokers, current electronic cigarettes (e-cigarette) users only, current conventional cigarettes (c-cigarette) users only, and dual users. OSA was assessed by three main signs and symptoms from the questionnaire. Multivariable logistic regression after adjusting for covariates was conducted to investigate the association of OSA with different smoking patterns.

The sample size was calculated as 233 to a population of 12,900000 with a margin error of 5% and a confidence interval of 95% using a sample size calculator.

**Results:** Among the 233 participants, the prevalence of OSA was higher among smokers compared to non-smokers.

the distribution of the prevalence of OSA among continuous smokers (16%), intermittent smokers (36%), and passive smokers (24%).

As for those undiagnosed with OSA, the percentage of smokers was 40%, passive smoking was 12.9%, and intermittent smokers were 17.1%.

As for OSA patients who were non-smokers, they constituted 24%.

By calculating the P-VALUE, which amounted to < 0.001, which indicates a relationship between smoking and sleep apnea.

**Conclusion:** Our study found that 68% of sleep apnea patients were smokers, with 24% passive smoking, 36% smoking regular cigarettes, 24% smoking hookah, and 8% smoking electronic cigarettes, which confirmed that sleep apnea is more related to smoking regular cigarettes than Electronic cigarettes

## INTRODUCTION

Obstructive sleep apnea syndrome (OSA) is a sleep-related breathing disorder characterised by snoring, repeated episodes of airflow cessation, hypoxemia during sleep, and daytime hypersomnolence caused by the collapse of the upper airway. Apnea attacks more than 15 times or apnea hypopnea index (AHI) >15 [2].

Moreover, the patients were accompanied by clinical symptoms such as snoring at night and sleepiness during the day.

OSA is related to a variety of diseases and complications, such as coronary disease, hypertension, diabetes,cerebral apoplexy, gastroesophageal reflux disease, orthodontic anomalies,and malignant tumour.[2]

They had common pathophysiology mechanisms, such as sympathetic activation, and systemic inflammation.

Moreover, the risk factors for OSA were obesity,sex,hyperglycemic,upper respiratory tract stenosis,hypertension,hyperlipidemia,Nasal obstruction, Acromegaly, Hypothyroidism,Alcohol,Congenital inferior position of the hyoid bone,advanced age,pregnancy,etc.. [2][3].

It is a known fact that smoking can lead to various respiratory diseases (chronic obstructive pulmonary disease,chronic bronchitis,chronic pulmonary heart disease,etc..),cardiovascular diseases (coronary heart disease,hypertension, etc..)and malignant tumour diseases (lung cancer,nasopharyngeal carcinoma,oral cancer,etc…) [7][8][9][10]

## Methods

### Study conduction

A link has been posted to a set of questions in the form of a questionnaire consisting of 28 questions, divided between 8 demographic questions, 10 about symptoms, 4 questions about smoking, and 8 sub-questions of the EPWORTH scale.

The study was cross-sectional, targeting a sample of Syrian society(233 participants), by publishing the link on social media and the Internet, according to a Google form.

### Statical study

The research results were studied using the statistical program Spss version 26.0, and any statistical p value equal to 0.05 or less was considered statistically significant.

The Chi-square law and t-test were used to study the relationship between the variables present in the study.

### Exclusion Criteria

● Patients above the age of 75 and less than 18 were excluded.
● Pregnant and breastfeeding women were excluded.
● Patients who were prescribed benzodiazepines,barbiturate were excluded.
● Patients diagnosed with COPD or heart failure.

### Inclusion Criteria

● Patients included were between the age of 18-75.
● Subjects with internet connection and offered their consent within the online form.

## Conclusion

The spread of breath-interruption during sleep in OSA was divided as 16% continuous smokers and 36%as intermittent smoking and 24% as negative smoking.so there is a relation between smoking and OSA.

## Recommendations

1- We noticed that 30% of the studied sample had not been diagnosed with OSA and had symptoms that intersect with OSA patients,which prompts us to recommend directing more attention to the disease and increasing community awareness of it, which helps in detecting and managing it in the best way and alleviating symptoms for patients, which may improve Quality of life for this group.

2- We found that 76% of OSA patients are exposed to smoking, which draws attention to the necessity of quitting smoking.

If this is not possible, we advise the patient to seek the assistance of a health authority to provide him with mechanisms and means to enable him to gradually quit smoking.

3- We recommend avoiding the use of sedatives, as we noted that 32% of OSA patients were taking sedatives.

4- We also recommend following a diet with the aim of losing weight, as we found that one of the factors that helps reduce the severity of sleep apnea is weight loss, as 13.3% of those who lost weight improved their sleep.

5- We noticed that the percentage of those who want to sleep during the day reached 72% of sleep apnea patients, which indicates the importance of getting adequate sleep to avoid annoying daytime symptoms of fatigue or lack of concentration, which helps prevent exposure to the risk of driving accidents.

It is a state of relaxation of the pharyngeal muscles during a period of deep sleep, characterized by episodes of narrowing and obstruction of the upper respiratory tract. This obstruction may cause:

1. Apnea stops: no air flow occurs, and in this case the obstruction becomes a complete obstruction.
2. Hypnea: A decrease in flow occurs as a result of narrowing of the airways.

➢ These attacks lead to fragmented sleep, meaning that the patient sleeps in intermittent periods, which leads to fatigue and lethargy during the next day.

Obstructive apnea (OSA) is considered a common disease, affecting about one billion people worldwide, and its prevalence in the United States of America is on average 27% of males and 13% of women. The percentage increases in Spain, Asia, and the black race. The probability of its development increases with age, and obesity plays an important role in its development, as its prevalence increases in obese people by a rate of 14% to 55%. [2]

### Symptoms

Obstructive apnea is characterized by frequent awakenings during sleep as a result of episodes of interrupted breathing, resulting in a group of daytime symptoms such as constant sleepiness, fatigue, and lack of concentration.

In addition to symptoms that the companion complains about: such as continuous night snoring and frequent tossing and turning during sleep, the companion may notice that the patient’s breathing stops several times during his sleep. [2]

The most important symptoms:

Excessive daytime sleepiness and fatigue.

Loud snoring in all sleep body positions.

A feeling of not getting enough sleep, even for long hours.

Difficulty concentrating and impaired cognitive function.

Decreased patient efficiency and performance at work.

Irritability and depression.

## Results

### Demographic characteristics of patients

The total number of patients, which amounted to 232, and their percentage according to gender were calculated, as the percentage of males was 52.4% and females were 47.6%.

Patients were also classified according to marital status. The majority were unmarried, who constituted 75% of the sample, while married people constituted 24% of them.

The sample was distributed between 52.8% students, 38.2% employees, and 6.5% unemployed.

The educational level of 76.8% of them was university, 53.5% specialized in the medical field, and 34.8% were outside the medical field. As shown in **Table 1**.

**Table (1).** demographic characteristics.

| Table (1) demographic characteristics |  |  |
| --- | --- | --- |
| 52.4% | MALE | gender |
| 47.6% | FEMALE |  |
| 24% | MARRIED | Marital status |
| 75% | SINGLE |  |
| 1.0% | WIDOW |  |
| 1.7% | NOT WORKER | job |
| 38.2% | EMPLOYER |  |
| 7.3% | RETIRED |  |
| 52.8% | STUDENT |  |
| 4.3% | SCHOOL | academic level |

| Table (1) demographic characteristics |  |
| --- | --- |
| 76.8% | UNIVERSITY |
| 18.9% | HIGHER |

**Table 2** shows the largest proportions of patients in Syrian cities, most of whom were from Damascus, as follows:

**Table-2.** the sample distribution in the Syrian governorates.

| Table-2 the sample distribution in the Syrian governorates |  |
| --- | --- |
| CITY | 82.4% |
| SUBURB | 17.6% |
| DAMASCUS | 42.5% |
| RIF-DIMASHQ | 13.7% |
| ALEPPO | 10.3% |
| LATTAKIA | 9.0% |

### Study of the relationship between smoking and obstructive sleep apnea (OSA) among a sample of Syrian society in terms of (type of smoking - duration - quantity)

**Table 3** shows the distribution of the prevalence of OSA among continuous smokers (16%), intermittent smokers (36%), and passive smokers (24%).

**Table 3.** The relationship between OSA and smoking.

| Table 3 - The relationship between OSA and smoking |  |  |  |  |  |
| --- | --- | --- | --- | --- | --- |
| SMOKING |  |  |  |  | OSA |
| TOTAL | INTERMITTENT | SMOKER | NEGATIVE SMOKING | NON SMOKER |  |
| (10.7%)25 | (36%)9 | (16%)4 | (24%)6 | (24%)6 | YES |
| (30%)70 | (17.1%)12 | (40%)28 | (12.9%)9 | (30%)21 | UNKNOWN |
| (59.2%)138 | (8.7%)12 | (28.3%)39 | (5.1%)7 | (58%)80 | NO |
| (100%)233 | (14.2%)33 | (30.5%)71 | (9.4%)22 | 45.9%)107<br>( | TOTAL |

As for OSA patients who were non-smokers, they constituted 24%.

While the relationship between the type of cigarettes used and the presence of OSA, we note in **Table 4** that 36% of OSA patients who s smoke regular cigarettes and 24% smoke hookah

**Table 4.** The relationship between the type of smoking used and OSA.

| Table 4 - The relationship between the type of smoking used and OSA |  |  |  |  |
| --- | --- | --- | --- | --- |
|  | OSA |  |  | TYPE OF SMOKING |
| total | no | unknown | yes |  |
| (51.5%)120 | (59.4%)82 | (42.9%)30 | (32%)8 | NON SMOKER |
| (26.2%)61 | (21%)29 | (32.9%)23 | (36%)9 | REGULAR |
| (5.6%)13 | (5.1%)7 | (8.6%)6 | 0 | MIX |
| (12%)28 | (9.4%)13 | (12.9%)9 | (24%)6 | hookah |
| (4.7%)11 | (5.1%)7 | (2.9%)2 | (5%)2 | E-CIGARETTE |
| (100%)233 | (59.2%)138 | (30%)70 | (10.7%)25 | TOTAL |

While 32.9% of those undiagnosed with OSA smoked regular cigarettes and 12.9% smoked hookah. Therefore, the type of smoking most commonly used by OSA patients was regular cigarette, followed by hookah

With the P-VALUE study, which was 0.049, there is a relationship between the type of smoking and OSA

The relationship between smoking duration and OSA is shown where we note that the percentage of undiagnosed OSA increases with continued smoking for more than 10 years, reaching 32.9%

The percentage of OSA patients who smoked for 0-5 years was 28% By studying the P-VALUE, which was 0.005, we find that there is a relationship between the duration of smoking and the presence of OSA

**Table 6** the relationship between snoring and OSA

**Table 5.** The relationship between osa and duration of smoking.

| Table 5 - The relationship between osa and duration of smoking |  |  |  |  |
| --- | --- | --- | --- | --- |
| total | OSA |  |  | duration of smoking |
|  | no | unknown | yes |  |
| (60.5%)141 | (65.0%)91 | (51.4%)36 | (56%)14 | Non smoker |
| (10.7%)25 | (8.7%)12 | (8.6%)6 | (28%)7 | 0-5 years |
| (8.6%)20 | (8.7%)12 | (7.1%)5 | (12%)3 | 5-10 years |
| (20.2%)47 | (16.7%)23 | (32.9%)23 | (4%)1 | >10 years |
| (100%)233 | (59.2%)138 | (30%)70 | (10.7%)25 | total |

**Table 6.** The relationship between OSA and snoring.

| Table 6 - The relationship between OSA and snoring |  |  |  |  |
| --- | --- | --- | --- | --- |
| TOTAL | OSA |  |  | SNORING |
|  | NO | UNKNOWN | YES |  |
| (67.8%)158 | (74.6%)103 | (58.6%)41 | (56%)14 | NO |
| (32.2%)75 | (25.4%)35 | (38.7%)29 | (44%)11 | YES |
| (100%)233 | (59.2%)138 | (30%)70 | (10.7%)25 | TOTAL |

We find that 44% of those with OSA suffer from snoring and 56% of them do not complain of snoring

The number of those complaining of snoring constituted 38.7% of those who were not diagnosed with OSA

The P-VALUE showed a relationship between snoring and OSA, reaching 0.026, which is statistically significant

Epworth scale

It is the internationally approved scale for the initial investigation of people with obstructive apnea. This scale is a set of 8 Questionnaire questions, and each question has 3 marks

We rely on the patient’s answer by placing a mark for each question, so that each question takes one of the numbers (3-2-1-0) and then we calculate the total (24-0). The higher the score, the more susceptible the patient is to contracting OSA

The scores were divided into areas to classify patients into three types as :follows

Normal sleepiness points (0-7)

points for moderate drowsiness (9-8)

Points for abnormal drowsiness (10-15)

Excessive drowsiness points (16-24)

**Table 7**

**Table 7.** The relationship between the EPWORTH scale and OSA.

| TOTAL | EPWORTH |  |  |  | OSA |
| --- | --- | --- | --- | --- | --- |
|  | EXCESSIVE<br>(16-24) | ABNORMAL<br>(10-15) | AVERAGE<br>(8-9) | NORMAL<br>(0-7) |  |
| 137 | 3 | 33 | 19 | 82 | NO |
| 70 | 5 | 22 | 28 | 15 | UNKNOWN |
| 25 | 5 | 8 | 3 | 9 | YES |
| 233 | 13 | 63 | 50 | 106 | TOTAL |

By applying the Epworth scale to the sample members, we found that patients with OSA were distributed among 36% of whom had normal sleepiness, 12% had moderate sleepiness, 32% had abnormal sleepiness, and 20% had excessive sleepiness

While 40% of those undiagnosed with OSA have symptoms of moderate sleepiness, 31% of them have abnormal sleepiness, and 7% have excessive sleepiness

The P VALUE was <0.001, and there is a statistically significant relationship between OSA and the Epworth scale

**Table 8** shows the relationship between age and sleep apnea

**Table 8.** : The relationship between age and OSA.

| Table 8: The relationship between age and OSA |  |  |  |  |
| --- | --- | --- | --- | --- |
| TOTAL | NO | UNKNOWN | YES | OSA |
| (9.9%) 23 | (10.1%) 14 | (8.6%) 6 | (12%) 3 | AGE<20 |
| (82.8%) 193 | (81.2%) 112 | (87.1%) 61 | (80%) 20 | AGE20-50 |
| (7.3%) 17 | (8.7%)12 | (4.3%) 3 | (8%) 2 | AGE>50 |
| (100%) 233 | (59.2%) 138 | (30%) 70 | (10.7%) 25 | TOTAL |

We note that 80% of OSA patients are between the ages of 20 and 50, while it was 87.1% among those not diagnosed with OSA

As for OSA patients who were over fifty, they constituted only 8%, and 12% were under twenty

In the study, P - VALUE = 0.78, and therefore there is no relationship between the two variables

**Table 9**

**Table 9:** The relationship between gender and OSA.

| Table 9: The relationship between gender and OSA |  |  |  |  |
| --- | --- | --- | --- | --- |
| TOTAL | OSA |  |  | GENDER |
|  | NO | UNKNOWN | YES |  |
| (52.4%)122 | (49.3%) 68 | (58.6%)41 | (52%) 13 | MALE |
| (47.6%)111 | (50.7%) 70 | (41.4%)29 | (48%)12 | FEMALE |
| (100%)233 | (59.2%)138 | (30%)70 | (10.7%)25 | TOTAL |

Shows the relationship between gender and sleep apnea

We note that 52% of OSA patients are male and 48% of them are female

While 58.6% of those undiagnosed with OSA were male and 41.4% of them were female

We found a P value = 0.447, and therefore there is no relationship between the two variables

**Table 10**

**Table 10.** Relationship between OSA and the field of university study.

| Table 10 Relationship between OSA and the field of university study |  |  |  |  |
| --- | --- | --- | --- | --- |
| total | no | unknown | yes | OSA |
| (37.8%) 88 | (31.9%)44 | (42.9%) 30 | (56%) 14 | Non medical |
| (51.5%)120 | (60.9%) 84 | (38.6%) 27 | (36%) 9 | Medicine |
| (% 4.3) 10 | (3.6%) 5 | (4.3%) 3 | (8%) 2 | pharmacology |
| (1.3%) 3 | 0 | (4.3%) 3 | 0 | nursery |
| (5.2%) 12 | (3.6%)5 | (10%) 7 | 0 | dentistry |
| 233 | (59.2%)138 | (30%) 70 | (10.7%) 25 | total |

In it, we discussed the relationship between the prevalence of sleep apnea and the field of medical and non-medical specialization

In comparison between the sample members from the medical field and individuals from outside medical specialties, we found that 15.9% of the individuals outside the medical field suffered from sleep apnea, while the percentage was lower among the sample members from the medical field, as it decreased by half in the field of human medicine, reaching 7.5% Through statistical study, we found that P-VALUE = 0.004, which is statistically significant, and from it there is a relationship between the two variables We found that 56% of those who complain of OSA are from outside the medical field, while 36% of them studied medicine and 8% studied pharmacy, while we did not find any individual whose specialty was dentistry or nursing

As for the relationship between the number of hours of sleep and obstructive sleep apnea (OSA), **Table 11** shows us We find that the percentage of those who sleep 7-8 hours constitutes 56% of those with OSA and constitutes about 25% of those who are not diagnosed The P value statistically reached < 0.01, from which we conclude that there is a relationship between them

**Table 11.** The relationship between OSA and the hours of sleep.

| Table 11 The relationship between OSA and the hours of sleep |  |  |  |  |
| --- | --- | --- | --- | --- |
| total | no | unknown | yes | OSA |
| (0.9%) 2 | 0 | (2.9%)2 | 0 | <4 sleeping hours |
| (19.7%)46 | (16.7%)23 | (30%)21 | (8%)2 | 5-6 sleeping hours |
| (36.9%)86 | (44.9%)62 | (31.4%)22 | (8%)2 | 6 -7 sleeping hours |
| (33%)77 | (32.6%)45 | (25.7%)18 | (56%)14 | 7-8 sleeping hours |
| (9.4%)22 | (5.8%)8 | (10%)7 | (28%)7 | >8 sleeping hours |
| (100%)233 | (59.2%)138 | (30%)70 | (10.7%)25 | total |

**Table 12**

**Table 12.**
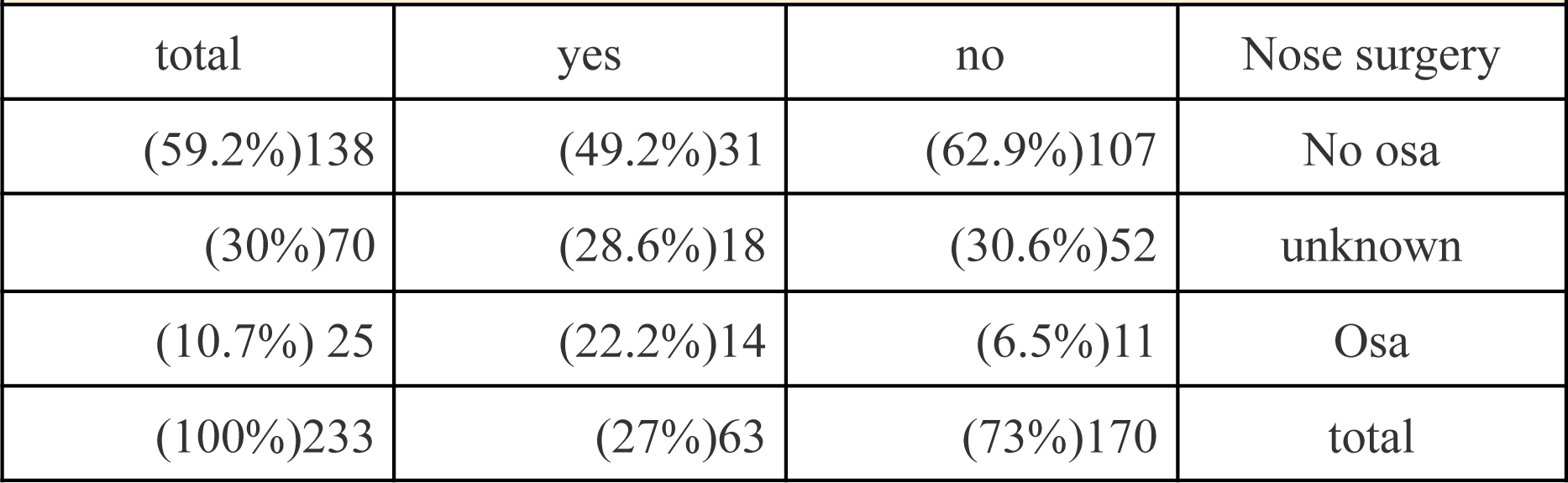
The relationship between nose surgery and OSA.

| Table 12 The relationship between nose surgery and OSA |  |  |  |
| --- | --- | --- | --- |
| total | yes | no | Nose surgery |
| (59.2%)138 | (49.2%)31 | (62.9%)107 | No osa |
| (30%)70 | (28.6%)18 | (30.6%)52 | unknown |
| (10.7%) 25 | (22.2%)14 | (6.5%)11 | Osa |
| (100%)233 | (27%)63 | (73%)170 | total |

While people who had nose operations had a higher percentage of sleep apnea than those who did not have operations, as 22.2% of operation patients suffered from OSA

The percentage decreased to 6.5% for the other group, and the relationship was accompanied by p value = 0.02, which was statistically significant

As for patients diagnosed with a rhythm deviation, we found that their rate of apnea increased more than three times compared to patients who did not suffer from a rhythm deviation, so the osa percentage for the first category reached 23%, compared to 6.5% for the latter, with a p value < 0.01, which is statistically significant..

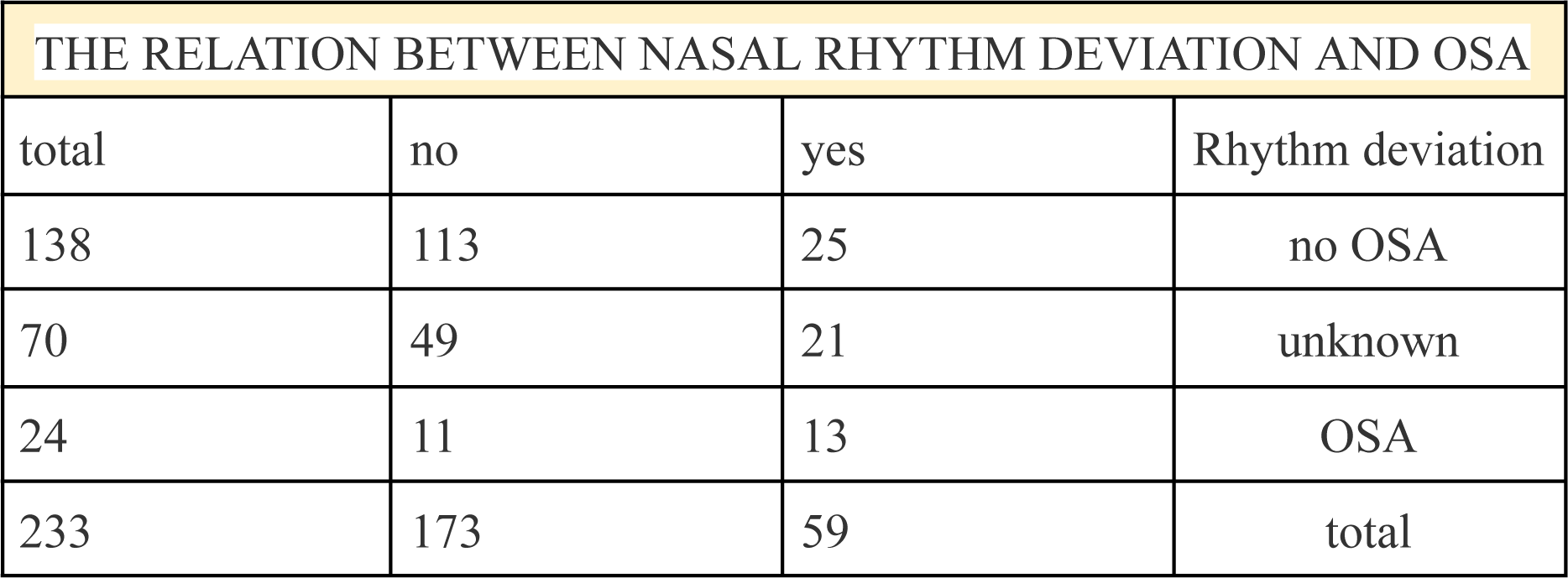

**Table 13**

**Table 13.** The relationship between having a positive family history and OSA.

|  |  |  |  |  |
| --- | --- | --- | --- | --- |
| total | no | unknown | yes | Family history |
| 138 | 70 | 47 | 21 | No OSA |
| 70 | 27 | 33 | 10 | unknown |
| 25 | 12 | 6 | 7 | OSA |
| 233 | 109 | 86 | 38 | total |

It studies the relationship between OSA and having a positive family history of OSA

The percentage of OSA patients with a positive family history is 28%, while the percentage of those with no family history of OSA is 48%, and 24% of them are unaware of the existence of a family history

According to P VALUE = 0.122, there is no relationship between them

**Table 14**

**Table 14.** Relationship between OSA and group of chronic diseases.

| Table 14 Relationship between OSA and group of chronic diseases |  |  |  |  |
| --- | --- | --- | --- | --- |
| total | no | unknown | yes | OSA |
| (87.1% ) 203 | (91.3%) 126 | (78.6%)55 | (88%) 22 | No chronic disease |
| (4.7%)11 | (2.2%)3 | (10%) 7 | (4%)1 | asthma |
| (2.6%) 6 | (3.6%) 5 | (1.4%) 1 | 0 | hypertension |
| (3%)7 | (1.4%) 2 | (% 7.1) 5 | 0 | diabetes |
| (2.6% ) 6 | (1.4%) 2 | (2.9%) 2 | (8%) 2 | COPD |
| (100%) 233 | (59.2%) 138 | (30%)70 | (10.7%) 25 | total |

It shows the relationship between OSA and a group of chronic diseases. We found that 78.6% of those undiagnosed with OSA do not suffer from a chronic disease, 10% of them suffer from asthma, and 7.1% have diabetes

We also noticed that 4% of OSA patients have asthma and 8% of them suffer from COPD

By studying P -VALUE = 0.022, thus there is a relationship between the two variables

**Table 15** shows the relationship between sleep apnea and BMI

**Table 15.** Relationship between BMI and OSA.

| Table 15 Relationship between BMI and OSA |  |  |  |  |
| --- | --- | --- | --- | --- |
| TOTAL | NO | UNKNOWN | YES | OSA |
| (19.7%) 46 | (18.8%)26 | (14.3%) 10 | (40%)10 | OVERWEIGHT |
| (80.3%)187 | (81.2%) 112 | (85.7%) 60 | (60%)15 | NORMAL WEIGHT |
| (100%)232 | (59.2%) 138 | (30%) 70 | (10.7%)25 | TOTAL |

We noticed that as patients gained weight, they were approximately two and a half times more likely to suffer from sleep apnea compared to patients with normal weight, as 22% of overweight patients were diagnosed with OSA By calculating P-VALUE = 0.02, it is statistically significant.

**Table 16**

**Table 16.** The relationship between alcohol consumption and OSA.

| Table 16 The relationship between alcohol consumption and OSA |  |  |  |  |
| --- | --- | --- | --- | --- |
| TOTAL | NO | UNKNOWN | YES | OSA |
| (92.3%)215 | (89.1%)123 | (97.1%) 68 | (96%)24 | NO ALCOHOL |
| (7.7%)18 | (10.9%)15 | (2.9%)2 | (4%)1 | ALCOHOL |
| (100%) 233 | (59.2%)138 | (30%)70 | (10.7%)25 | TOTAL |

Shows the relationship between OSA and the patient’s alcohol intake We found that 96% of OSA patients do not drink alcohol and only 4% do. For those undiagnosed with OSA, we found that 97.1% of them do not drink alcohol.

By calculating P -VALUE = 0.094, therefore there is no relationship between the two variables..

**Table 17**

**Table 17:** The relationship between taking sedative medications and OSA.

|  |  |  |  |  |
| --- | --- | --- | --- | --- |
| TOTAL | NO | UNKNOWN | YES | OSA |
| (91%)212 | (93.5%)129 | (94.3%)66 | (68%)17 | NO SEDATIVE MEDICATIONS |
| (9%)21 | (6.5%)9 | (5.7%)4 | (32%)8 | SEDATIVE MEDICATIONS |
| (100%) 233 | (59.2%)138 | (30%)70 | (10.7%)25 | TOTAL |

Shows the relationship between OSA and taking sedative medications

We find that 68% of OSA patients do not use sedative medication, and 32% of them use sedative medication

Among those who were not diagnosed with OSA, the percentage was 5.7% of them taking sedative medication

By calculating P-VALUE = 0.01, there is therefore a relationship between the two variables

**Table 18**

**Table 18.**
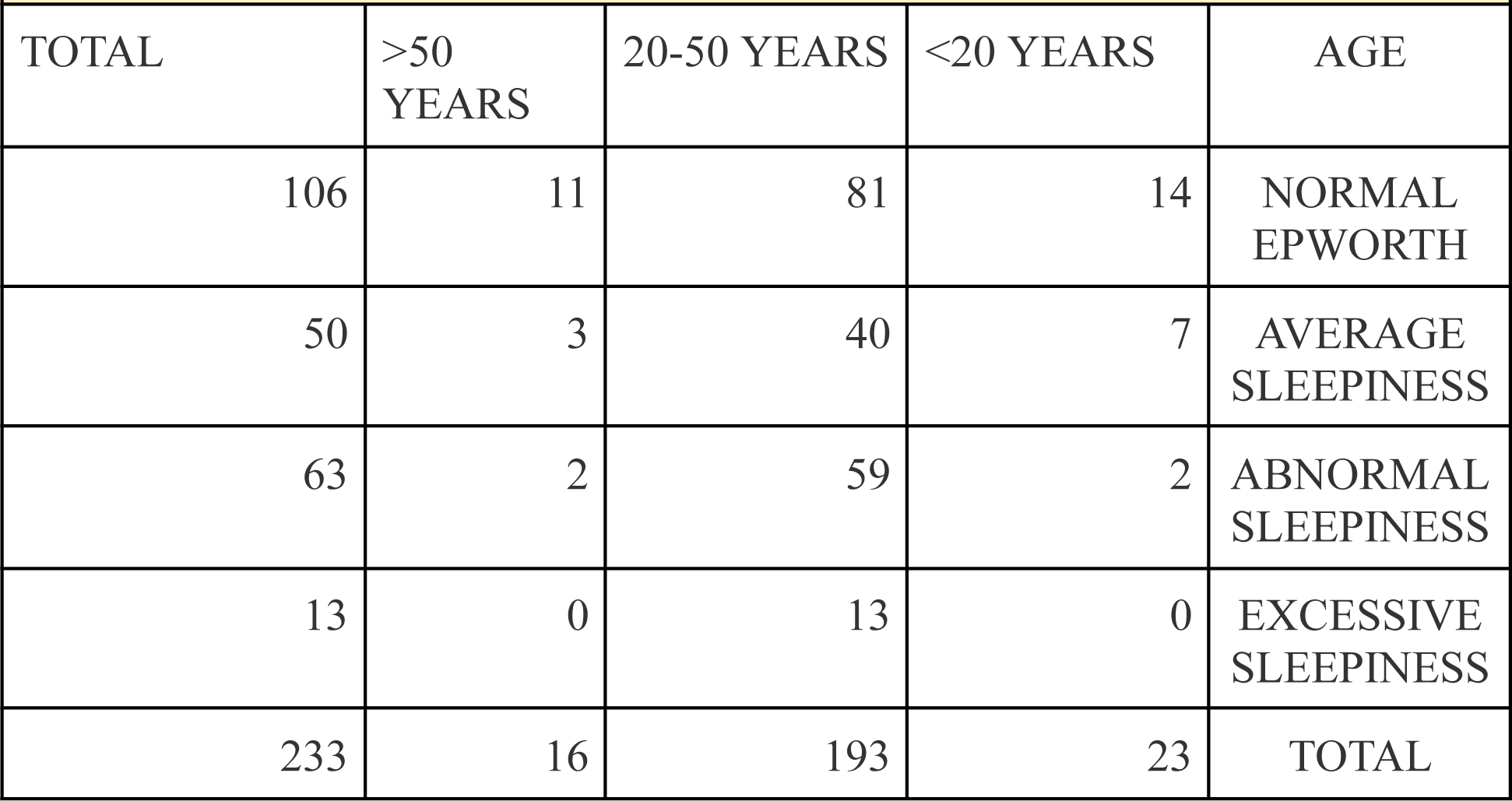
Relationship between age and Epworth scale.

It studies the relationship between age and the results of applying the Epworth scale

It shows that 58.03% of those aged between 20-50 years had an Epworth scale above the normal limit (moderate sleepiness - abnormal sleepiness - excessive sleepiness), while the percentage decreased to about half, 31.2%, among those aged over fifty

As for those aged < 20 years, the percentage was 39.1%, and by calculating P - VALUE = 0.058, we find that there is no relationship between age and obtaining Epworth results above the normal limit

**Table 19**

**Table 19.** The relationship between smoking and the Epworth scale.

| Table 19 The relationship between smoking and the Epworth scale |  |  |  |  |  |
| --- | --- | --- | --- | --- | --- |
| total | Non smoker | Negative smoking | smoker | interruptally | Smoking |
| 106 | 57 | 10 | 30 | 9 | Normal epworth |
| 50 | 21 | 8 | 14 | 7 | Average sleepiness |
| 63 | 21 | 4 | 23 | 15 | Abnormal sleepiness |
| 13 | 7 | 0 | 4 | 2 | Excessive sleepiness |
| 233 | 106 | 22 | 71 | 33 | Total |

Shows the relationship between smoking and Epworth scale results

It appears that the percentage of those exposed to all types of smoking and having an Epworth above the normal limit was 33.04%, and therefore, according to P VALUE = 0.075, there is no statistical relationship between the two variables

**Table 20**

**Table 20.** The relationship between OSA and the need for hospitalization.

|  |  |  |  |  |
| --- | --- | --- | --- | --- |
| TOTAL | NO | UNKNOWN | YES | OSA |
| 193 | 112 | 59 | 22 | NO<br>HOSPITALISATION |
| 3 | 1 | 0 | 2 | HOSPITALISATION |
| 196 | 113 | 59 | 24 | TOTAL |

Shows the relationship between OSA and the need for hospitalization and oximetry

We note that the percentage of OSA patients who did not require hospitalization was 91.6%, and therefore there is no relationship between the disease and the need for hospitalization

## DISCUSSION

Our study found that 68% of sleep apnea patients were smokers, with 24% passive smoking, 36% smoking regular cigarettes, 24% smoking hookah, and 8% smoking electronic cigarettes, which confirmed that sleep apnea is more related to smoking regular cigarettes than Electronic cigarettes, as this is consistent with an American study that appeared last year studying the relationship between electronic and regular cigarette smoking and sleep apnea. The percentage of OSA patients was male, 52%, while female, it was 48%. The percentage in the studied sample was close, and there is no relationship between gender and the disease, contrary to what is known and confirmed by the American study.

By comparing the sample members from the medical field and individuals from outside medical specialties, we found that 15.9% of the individuals outside the medical field suffered from sleep apnea, while the percentage was lower among the sample members from the medical field, which decreased by half in the field of human medicine, where it reached 7.5%.

This contradicts the American study, which did not find a relationship between the disease and the level of education.

Patients with sleep apnea with a history of asthma constituted 4%. As for the prevalence of snoring among OSA patients, it reached 44%, distributed between 16% who snore daily and 24% of whom snore once or twice a week. In contrast, the percentage of absence of snoring in the sample of sleep apnea patients was 56%.

By studying the marital status of our sample members, we found that 92% of sleep apnea patients were single and only 8% were married, unlike the American study.

We also noticed that 40% of those suffering from sleep apnea had an obesity rate that exceeded the normal limit, and the American study confirmed that BMI is a factor affecting OSA..

Also, the fact that 96% of OSA patients do not drink alcohol negates the relationship between drinking alcohol and sleep apnea, unlike the other study that found a relationship between them.

By applying the Epworth scale to the sample, we found that 36% of OSA patients had a normal Epworth

12% had moderate sleepiness, 32% had abnormal sleepiness, and 20% had excessive sleepiness.

Statistics were also conducted on the prevalence of aplasia in OSA patients, which constituted 20%, and the P-VALUE was = 0.017, which is statistically significant.

We also studied the relationship between snoring and Epworth values above the normal limit, and we found that 64.2% of those who had Epworth values above the normal limit suffered from snoring, and the P-VALUE study reached 0.026, which is statistically significant, confirming the existence of a relationship between the two variables.

When we asked about behaviours followed by the sample members that improved their condition, 13.3% of them responded by losing weight, 5.2% by having nose surgery, 4.7% by stopping smoking, 9.9% by changing the sleeping position, 3% by removing adenoids, and 3.4% by using sleeping medications.

## Data Availability

all data are available at request

## CONFLICT OF INTEREST

The authors declare that there is no conflict of interest.

## Funding

there was no funding for this study.

## Notes

### Competing Interest Statement

The authors have declared no competing interest.

### Author Declarations

Ethics committee of Syrian Private University gave approval for this work

